# Large Language Models Improve Cancer Survival Prediction Using Real-World Clinical Notes

**DOI:** 10.1101/2025.08.17.25333835

**Authors:** Niklas Kiermeyer, Tim Lenfers, Amin Dada, Julian Friedrich, Sameh Khattab, Eric Knop, Jan Egger, Markus Pauly, Andreas Jung, Grégoire Montavon, Jens T. Siveke, Marcel Wiesweg, Stefan Kasper, Ulf P. Neumann, Frederick Klauschen, Sylvia Hartmann, Martin Schuler, Philipp Keyl, Jens Kleesiek, Julius Keyl

## Abstract

In medical documentation, vast amounts of unstructured text are generated that are still underutilized in current prognostic models. We investigate the potential of self-hosted large language models (LLM) to extract clinically meaningful, patient-specific information from routine clinical notes for personalized risk stratification in cancer care.

We collected real-world medical notes from 2,708 non-small cell lung cancer (NSCLC) patients and 814 colon cancer patients documented before treatment at a large comprehensive cancer center. LLMs extracted key prognostic indicators, including comorbidities, metastatic sites, and qualitative descriptors of patient condition, in a zero-shot manner without prior task-specific training. Integrating these LLM-derived features into machine learning models significantly improved the prediction of overall survival compared to TNM staging alone (C-Index: NSCLC, 0.72 vs 0.64; colon cancer, 0.70 vs 0.59), and surpassed models using text embeddings. Based on the LLM-informed risk scores, patients were stratified into four distinct risk groups, enabling reclassification of 61.4% of NSCLC and 68.3% of colon cancer patients. Analysis of model drivers revealed that LLM-derived factors, such as the physical condition, substantially modulated the prognostic impact of TNM stage.

These findings highlight the potential of self-hosted LLM to extract clinically meaningful information from unstructured clinical documentation and support clinical decision-making.

## Introduction

Unstructured medical reports contain a wealth of doctor-curated patient-specific information that is currently not fully exploited for patient stratification. Especially host-specific information, such as the patient’s physical condition and disease-related symptoms, has substantial influence on patient outcomes and should be an integral part of the clinical decision process and cancer research, complementing tumor-specific information. As these data are currently often not assessed in a structured format, clinical notes remain the only source that regularly incorporates this information. However, as current prognostic systems rely on structured data, this data source remains largely untapped in deployed prognostic systems. Even current multimodal models, which aim to incorporate comprehensive patient information and show great promise in cancer treatment guidance and biomarker discovery, mostly neglect the wealth of information hidden in text data.^1–7^ This is due to the fact that extracting relevant information from unstructured text was traditionally a labor-intensive and error-prone process, which often required domain experts to manually review and correct the results. Yet, recent advances in natural language processing (NLP) and Large Language Models (LLMs) have largely removed this barrier, enabling the automated extraction of meaningful information from unstructured text data.^8–10^ In particular, the rapid pace of research around open-source LLMs such as Meta’s Llama has created an environment where new and improved models are now released almost monthly.^11,12^ Moreover, it enables hospitals to self-host these LLMs, which is essential for use in routine clinical practice and in the context of data privacy. However, the clinical implementation of these existing approaches remains limited, and most approaches are applied on small, non-real-world datasets limiting their generalizability.^13,14^

Here, we developed an explainable, end-to-end survival-prediction framework to integrate LLM-extracted features and specialized text embeddings on large, real-world cancer cohorts. To this end, we employed Llama 4 Scout and a dedicated embedding model to process EHR notes and discharge reports from 2,708 NSCLC patients and 814 colon cancer patients. Our study shows the potential of LLMs to support personalized risk stratification by extracting patient-specific information from unstructured clinical text.

## Results

### Cohort description

Our real-world dataset comprised 2,708 patients diagnosed with non-small cell lung cancer (NSCLC) and a validation cohort of 814 patients with colon cancer treated at the University Hospital Essen (NSCLC: 2017-2025, colon cancer: 2010-2025, **Fig.1 + Tab. 1**). For all included patients, unstructured text data from EHR notes and discharge reports prior to the start of cancer therapy were collected. In the NSCLC cohort (median age 67.3 years; 41% female), 996 patients experienced the event (death), while 1712 were right-censored. Adenocarcinoma was the most common histology (61.5%), followed by squamous cell carcinoma (29.8%), large cell carcinoma (4.5%), and adenosquamous carcinoma (4.2%). Stage IV disease accounted for 48.6% of cases (Stage I 23.1%, Stage III 19.0%, Stage II 9.4%). In the colon cancer cohort (median age 66.7 years; 48% female), 510 patients experienced the event, and 304 were right-censored. Stage IV disease was present in 57.7% of cases (Stage III 16.6%, Stage II 14.7%, Stage I 11.3%).

### Structured extraction of metastatic patterns and comorbidities using LLM

To assess whether unstructured text from EHR notes and discharge reports could improve patient stratification beyond TNM staging, we used Llama 4 Scout to automatically extract clinically relevant features prior to the start of treatment at our center. The extracted features comprised metastatic sites, comorbidities (encoded as ICD codes), and seven LLM-inferred patient condition indicators (PCIs): mobility impairment, pain, B-symptoms, high-risk status, dyspnoea, complicated disease course, and abnormal physical examination. In addition, the LLM generated two composite scores for each patient: a physical condition score and a survival score inferred only from EHR notes and discharge reports. We observed distinct metastasis patterns in the stage 4 patients of our two cohorts. In NSCLC patients, the most frequently LLM-extracted metastatic sites were the lungs (25%), lymph nodes (17%), and bones (13%) (**Fig. 2A**). In contrast, LLM-extractions for colon cancer patients included predominantly liver metastases (40%), followed by lymph nodes and lung metastases. Extraction quality of metastases was validated on a subset of stage IV patients for which structured information was available in our database (NSCLC: 968, colon cancer: 404; **Supp. Fig. 1**).

**Figure 1:**
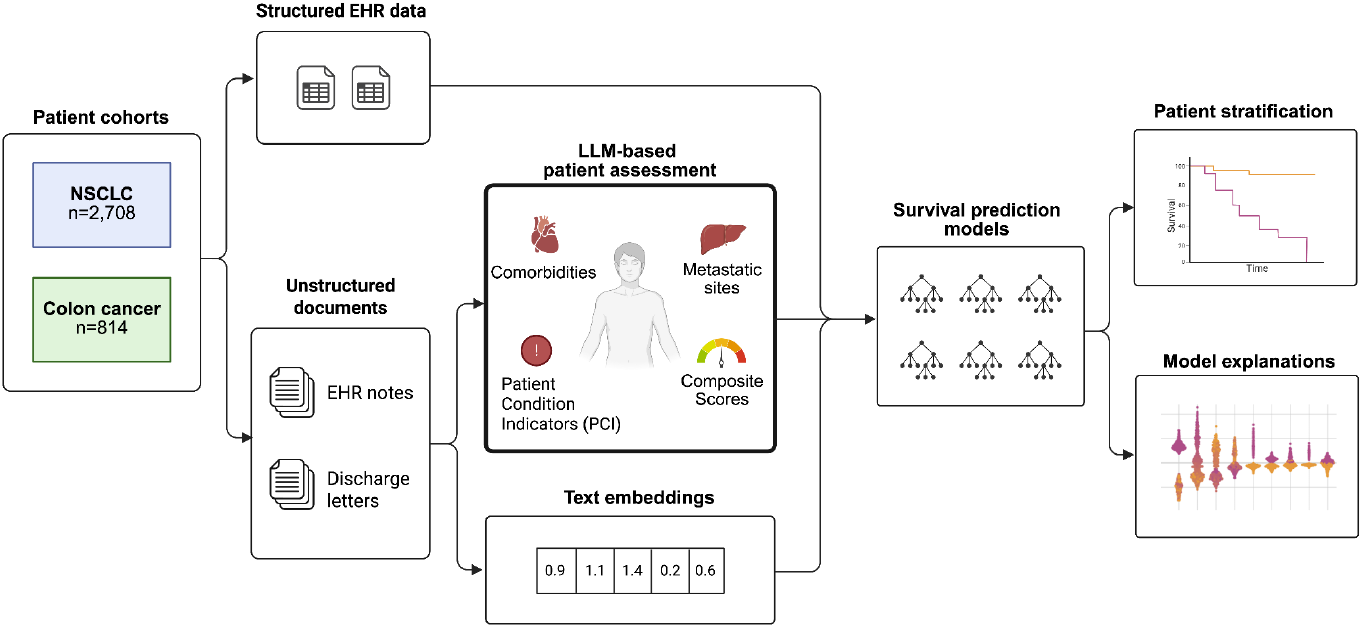
Study workflow.

**Figure 2:**
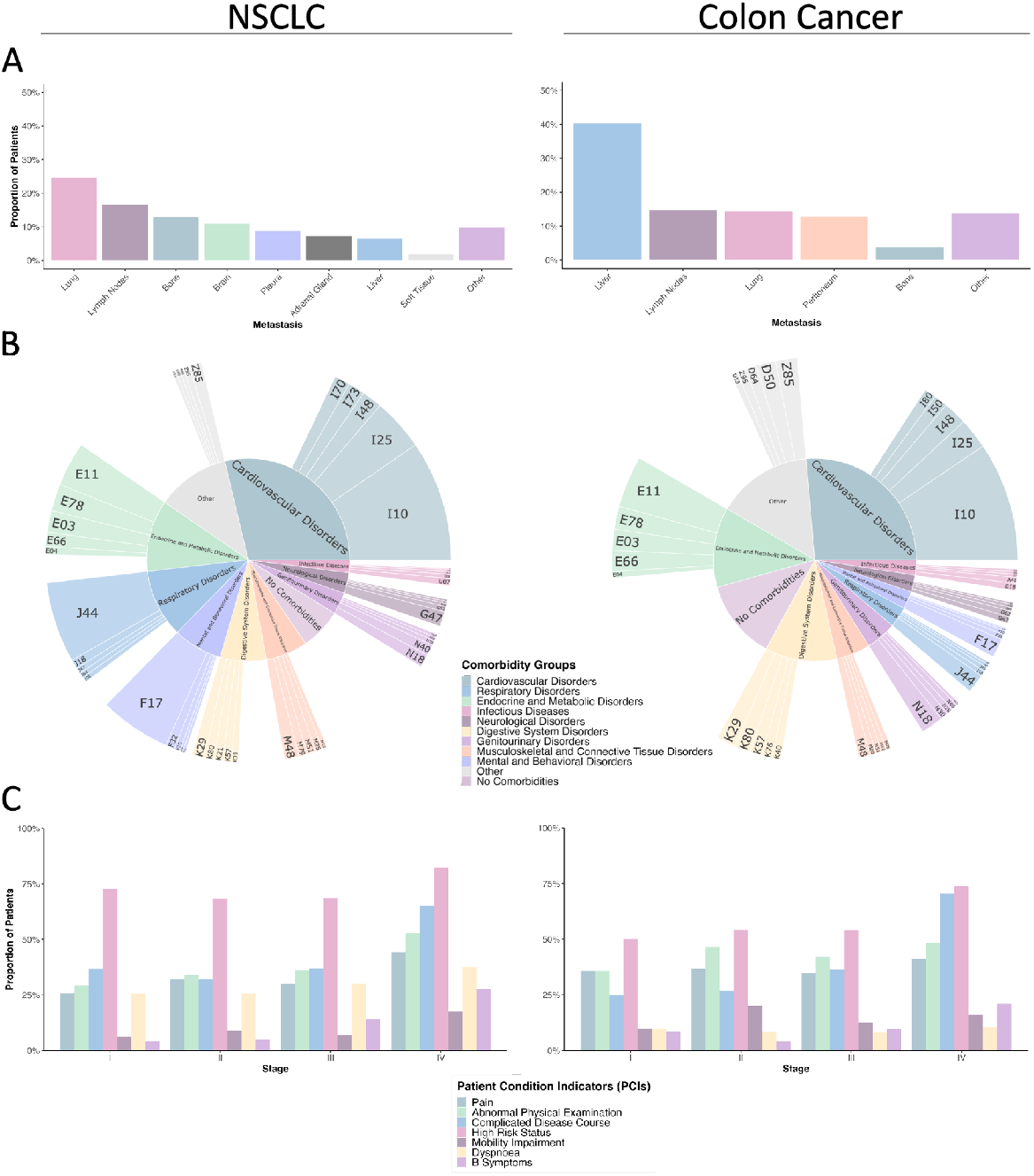
LLM-extracted features in NSCLC and colon cancer cohorts. **A:** Distribution of LLM-extracted metastatic sites in stage 4 NSCLC (left) and colon cancer (right) patients. **B:** Distribution of five most frequent comorbidities in NSCLC (left) and colon cancer (right) patients. **C:** Proportion of patients with a positive (True) binary LLM-extracted feature.

The LLM-extracted comorbidities also showed cancer-specific patterns after grouping by organ system. As expected, respiratory comorbidities were more frequently extracted in the NSCLC cohort, while disorders of the digestive system were more common in colon cancer patients. However, there were also similarities between both cohorts, as Cardiovascular diseases emerged as the most prevalent comorbidity in both cohorts. The top three codes extracted were I10 (hypertension), I25 (ischemic heart disease), and I48 (atrial fibrillation, **Fig. 2B**).

### LLM derive clinically relevant patient condition indicators

Next, we assessed the distribution of LLM-inferred PCI across cohorts and UICC stages. The proportion of patients with a high-risk status increased with disease stage in both cancers, rising from 73% at stage I to 82% by stage IV in NSCLC, and from 50% to 74% in colon cancer (**Fig. 2C**). Similarly, the prevalence of patients with complicated disease courses and abnormal findings on physical examination increased with advancing stages, reflecting higher disease burden and clinical severity at later stages.

Mobility impairment showed a markedly different pattern between cohorts: In NSCLC patients, it was uncommon in early-stage disease (<9% at stages I-II) but increased to 17% by stage IV. Colon cancer patients exhibited mobility impairment of 10-20% in stages I-II, but this did not increase by stage IV (16%). Pain prevalence in NSCLC increased from 26% at stage I to 44% at stage IV, paralleled by a rise in dyspnoea (from 26% to 37%). Conversely, colon cancer patients had relatively stable pain (35-41%) and low dyspnoea rates (<10%) regardless of stage. B-symptoms were rare (<10%) across stages I-II in both cohorts, but surged in stage IV (28% in NSCLC; 21% in colon cancer), suggesting systemic involvement in advanced disease.

### Prognostic impact of LLM-derived patient condition indicators

To assess the prognostic impact of the LLM-derived PCIs, we performed univariate cox regression analyses for descriptive purposes, followed by multivariate Cox models to account for potential confounders (**Table 2 + Supp. Table 1**). In univariate analysis of the NSCLC cohort, advanced tumor stage, male sex, and all PCIs were significantly associated with worse overall survival (OS). Among the PCI, mobility impairment was the strongest predictor of mortality (HR 2.64, p<0.001), followed by complicated disease courses (HR 2.19, p<0.001) and abnormal physical examination (HR 2.01, p<0.001). In multivariate analysis, high-risk status, abnormal physical examination, complicated disease course, and mobility impairment remained significant. However, high-risk status was associated with improved OS in multivariate analysis (HR 0.71, p<0.001).

**Table 1:** Characteristics of the patient cohorts. Bracketed values represent median overall survival (OS) time in months and the 95% confidence intervals (CIs).

|  | NSCLC<br>(n=2,708) | Colon cancer<br>(n=814) |
| --- | --- | --- |
| **Median Age** | 67 (Range: 24-91) | 66.0 (Range: 22-97) |
| **Sex** |  |  |
| Female | 1,135 (41.9%) | 392 (48.2%) |
| Male | 1,573 (58.1%) | 422 (51.8%) |
| **Stage** |  |  |
| I | 625 (23.1%) [not reached] | 92 (11.3%) [not reached] |
| II | 254 (9.4%) [not reached] | 120 (14.7%) [not reached] |
| III | 514 (19.0%)<br>[40.70, 95% CI: 27.62–51.72] | 135 (16.6%) [not reached] |
| IV | 1,315 (48.6%)<br>[18.90, 95% CI: 16.87–21.67] | 467 (57.4%)<br>[25.05, 95% CI: 20.94–32.38] |
| **NSCLC Histology** |  |  |
| Adenocarcinoma | 1,666 (61.5%) | - |
| Squamous Cell Carcinoma | 807 (29.8%) | - |
| Large Cell Carcinoma | 122 (4.5%) | - |
| Adenosquamous Carcinoma | 113 (4.2%) | - |
| **Text availability** |  |  |
| EHR notes | 2,630 (97.1 %) | 788 (96.8 %) |
| Discharge reports | 1,946 (71.9 %) | 474 (58.2 %) |

**Table 2:** NSCLC cohort results of univariate and multivariate Cox proportional-hazards models evaluating the association between LLM-extracted covariates and overall survival. Structured EHR data comprises all fields originally available in structured format, whereas LLM-inferred variables are those derived by the model from unstructured medical documentation.

|  | Univariate analysis |  | Multivariate analysis |  |
| --- | --- | --- | --- | --- |
| Structured EHR Data | HR (95% CI) | P value | HR (95% CI) | P value |
| Age at Treatment (per 1 SD) | 1.03 (0.97-1.1) | 0.301 | 1.11 (1.04-1.18) | **0.001** |
| Stage II vs I | 1.53 (1.08-2.15) | **0.016** | 1.47 (1.04-2.08) | **0.028** |
| Stage III vs I | 2.34 (1.80-3.04) | **<0.001** | 2.37 (1.82-3.09) | **<0.001** |
| Stage IV vs I | 4.10 (3.26-5.14) | **<0.001** | 3.62 (2.86-4.57) | **<0.001** |
| Sex (male) | 1.34 (1.18-1.53) | **<0.001** | 1.36 (1.19-1.55) | **<0.001** |
| Adenosquamous vs Adenocarcinoma | 1.09 (0.8-1.47) | 0.599 | 1.2 (0.88-1.63) | 0.253 |
| Large Cell vs Adenocarcinoma | 1.05 (0.77-1.43) | 0.767 | 1.16 (0.84-1.58) | 0.368 |
| Squamous cell vs Adenocarcinoma | 1.07 (0.93-1.23) | 0.354 | 1.06 (0.88-1.63) | 0.443 |
| **LLM-Inferred Variables** |  |  |  |  |
| High-Risk Status | 1.41 (1.21-1.64) | **<0.001** | 0.71 (0.59-0.86) | **<0.001** |
| Abnormal Physical Examination | 2.01 (1.78-2.28) | **<0.001** | 1.31 (1.11-1.54) | **<0.001** |
| Dyspnoea | 1.56 (1.38-1.77) | **<0.001** | 1.07 (0.92-1.24) | 0.380 |
| Complicated Disease Course | 2.19 (1.92-2.5) | **<0.001** | 1.63 (1.37-1.94) | **<0.001** |
| B-symptoms | 1.99 (1.72-2.3) | **<0.001** | 1.02 (0.86-1.2) | 0.821 |
| Pain | 1.62 (1.43-1.84) | **<0.001** | 1.07 (0.93-1.24) | 0.335 |
| Mobility Impairment | 2.64 (2.25-3.09) | **<0.001** | 1.78 (1.49-2.12) | **<0.001** |

In the colon cancer cohort, all of the LLM-derived PCIs were significant univariate predictors (**Supp. Table 1**). Of these, mobility impairment had the strongest effect on OS (HR 2.4, p<0.001), followed by B-symptoms (HR 2.11, p<0.001) and a complicated disease course (HR 1.92, p<0.001). In multivariate analysis, only mobility impairment (HR 1.65, p=0.005) and B-Symptoms (HR 1.4, p=0.025) remained significant among the PCI.

### LLM-derived composite scores for physical condition and survival

To enable the LLM to assess patients more comprehensively beyond closely defined markers, we prompted it for two composite scores, a physical condition score (0-100) and a survival score (0-100), based on each patient’s discharge report and EHR notes, resulting in up to four scores per patient. Higher values denote better physical status and greater likelihood of survival. The histograms showed that both physical condition and survival scores are not distributed uniformly for NSCLC and colon cancer, with certain score ranges being notably overrepresented (**Supp. Fig. 2**). In addition, a correlation was observed between both scores and patient condition indicators (**Supp. Fig. 3**). In univariate analysis, both LLM-derived composite scores were significantly associated with OS in the NSCLC cohort (physical condition score: HR 0.82, 95% CI: 0.78-0.86, p<0.001, survival score: HR 0.81, 95% CI: 0.77-0.86; p < 0.001; **Supp. Table 2**). In multivariate analysis, the physical condition score retained independent prognostic value (p<0.001).

In univariate analysis of the colon cancer cohort, both the LLM-inferred physical condition score (HR 0.79, 95% CI: 0.72-0.86, p<0.001) and survival score (HR 0.69, 95% CI: 0.62-0.77, p<0.001) were significantly associated with OS. Both scores remained independent prognostic markers in multivariate analysis (physical condition: p=0.02, survival: p=0.01, **Supp. Table 3**).

Binarizing each LLM-inferred score at its median into “low” and “high” groups yielded a clear stage-wise separation in the NSCLC cohort (**Fig. 3**): all log-rank tests were significant (p<0.05) except the survival score in stage III and the physical condition score in stage I. The colon cancer cohort showed the same overall pattern, though smaller sample sizes in stages I-III limited statistical power (**Supp. Fig. 4**).

**Figure 3:**
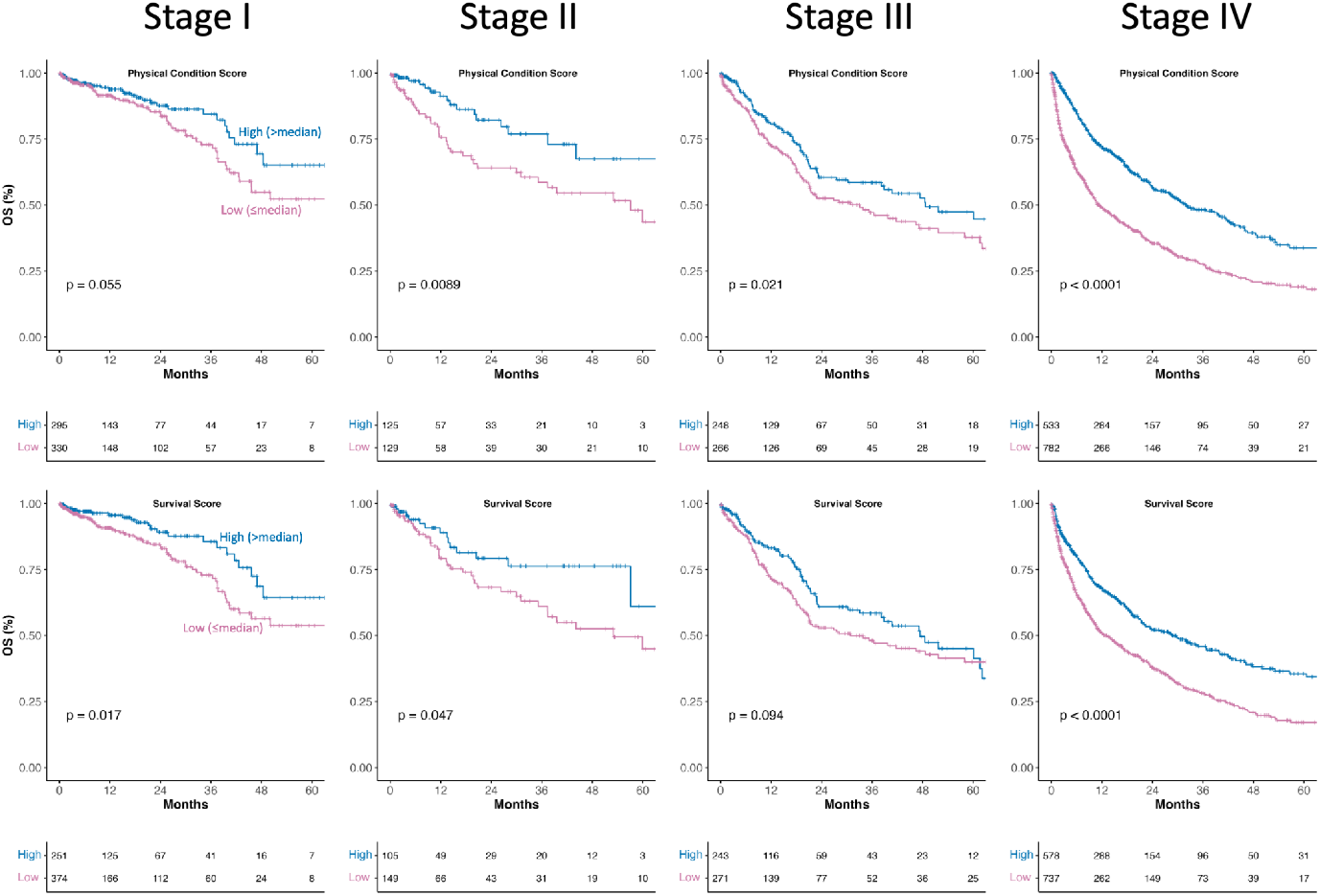
Kaplan-Meier survival curves for NSCLC patients stratified by LLM-inferred physical condition and survival scores at disease stages I–IV. Patients were dichotomized by the median within their respective stage into high-score (> median; blue) and low-score (≤ median; magenta) groups. Survival differences between groups were assessed by the log-rank test, and corresponding p-values are indicated in each panel.

### Comparing LLM-derived information and text embeddings for survival prediction

Next, we investigated whether unstructured EHR notes could improve survival prediction beyond tumor staging. The text information was used in two different forms: (1) structured features derived by LLM and (2) raw text embeddings generated by a specialized embedding model.^15,16^ We trained Random Survival Forest (RSF) models and compared four results: Tumor staging alone, RSF trained on baseline clinical variables (stage, age, sex), RSF trained on baseline plus text embeddings, and RSF trained on baseline plus LLM-derived features.^17,18^ Performance was evaluated using ten-fold cross-validated C-indices, with statistical significance determined by a paired Wilcoxon signed-rank test. In the NSCLC cohort, the text-embedding model significantly outperformed TNM staging (0.69 vs. 0.64, p=0.002, **Fig. 4A**) and the baseline model (0.69 vs. 0.64, p=0.002). The LLM-extractions model further improved median C-Index significantly over staging alone (0.72 vs. 0.64, p=0.002), versus the baseline model (0.72 vs 0.64, p=0.002) and versus the text-embedding model (0.72 vs 0.69, p=0.014).

**Figure 4:**
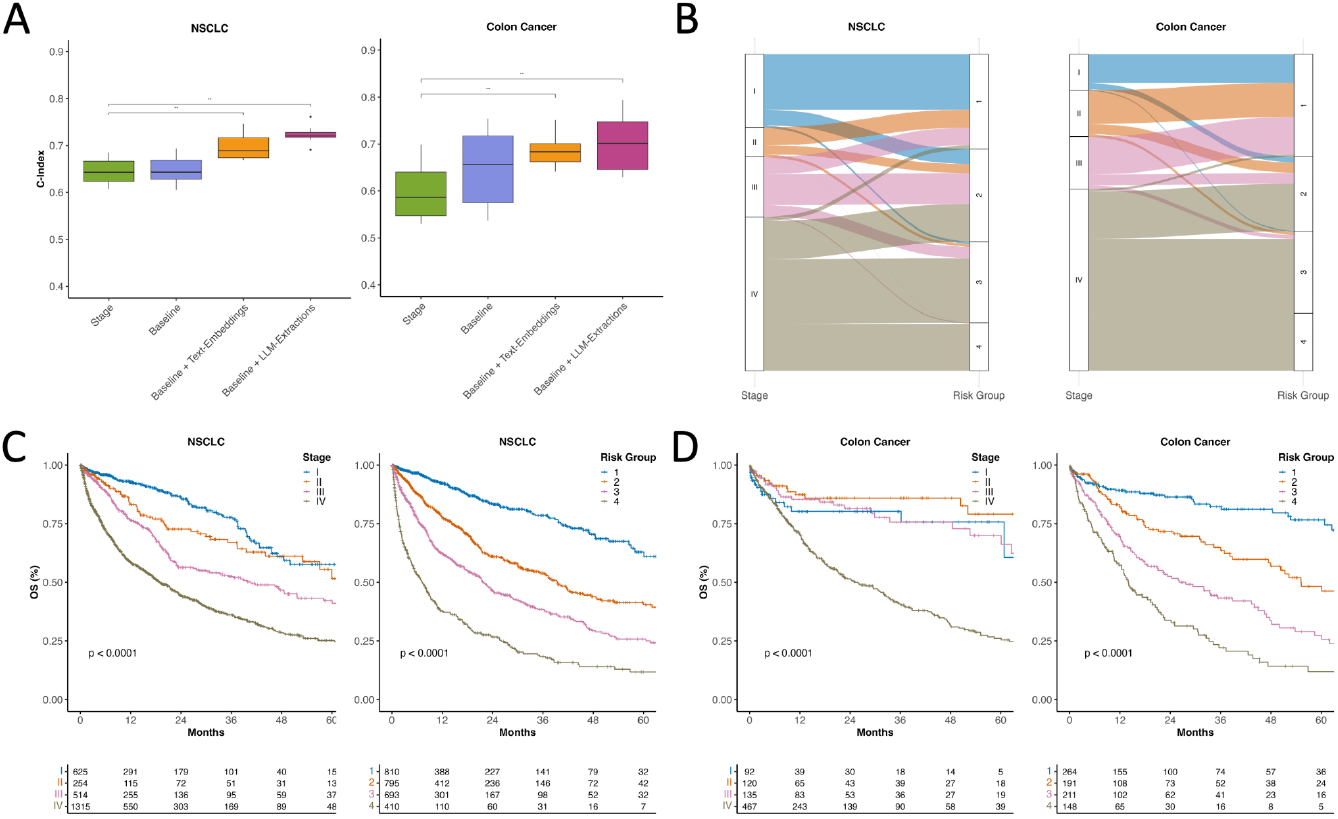
Prognostic Impact of baseline versus LLM-extracted features in NSCLC and colon cancer. **A:** Random Survival Forest results on a 10-fold CV on text embedding and LLM-derived information compared to a baseline model and tumor staging alone. All pairwise comparisons use the paired Wilcoxon signed-rank test. **B:** Sankey diagrams of patient re-stratification from clinical stage to RSF-predicted risk group in NSCLC and colon cancer, using an RSF trained on the combined baseline data and LLM-extracted features. **C:** Kaplan-Meier curves for NSCLC stratified by stage (left) and RSF-generated risk groups (right). **D:** Kaplan-Meier curves for colon cancer stratified by stage (left) and RSF-generated risk groups (right).

These results could be validated in the colon cancer cohort, where the text-embedding model reached a median C-Index of 0.68, outperforming stage (0.59, p=0.002) and baseline model (0.66, p=0.193). The LLM-derived model further improved prediction results compared to text embeddings (0.70 vs 0.68, p=0.275).

### LLM-enhanced prediction model enables improved patient stratification

Using predictions from the RSF model trained on the baseline data combined with the LLM-derived information, we stratified patients into four distinct risk groups according to their predicted cumulative hazards (see Methods, **Fig. 4B**). Comparing the documented tumor stages and the risk groups predicted by our model, we observed that in early-stage patients (I-II), our model reclassified patients only modestly into adjacent risk groups. In contrast, patients with more advanced stages (III-IV) were distributed across all four risk groups. This pattern was largely similar in NSCLC and colon cancer patients.

In Kaplan-Meier analysis, survival curves for both NSCLC and colon cancer cohorts showed more distinct separation between risk groups defined by our machine learning model with LLM-extracted features than between TNM stages (log-rank p<0.001; **Fig. 4C, D**). This enhanced stratification was particularly pronounced among early-stage patients in both cohorts. The greatest absolute impact of the LLM-enhanced model, however, was observed in stage IV, which represented the largest subgroup in both cohorts. Within this group, the model identified substantial prognostic heterogeneity, reclassifying the majority of patients into lower risk groups (NSCLC: 915 out of 1315, 69.6%, colon cancer: 319 out of 467, 68.3%). These findings underscore the potential of LLM-derived unstructured information to uncover prognostic heterogeneity not captured by UICC staging alone.

### LLM-derived features influence model predictions

To understand the decision-making process of the LLM-enriched prediction model, we conducted a feature importance analysis using Shapley Additive Explanations (SHAP, **Fig. 5A**).^19^ In both of our cancer cohorts, tumor stage was identified as the most influential variable: Among LLM-derived features, Complicated Disease Course ranked second in NSCLC. The LLM-inferred Survival and Physical Condition scores (third and fourth in importance). Abnormal physical examination, mobility impairment, and older age further increased risk. As expected, metastatic involvement of the liver, bone, or brain also increased predicted risk. The colon cancer model generally showed a similar order of feature importance (Spearman’s ρ = 0.758, p<0.001). After stage and age, the LLM-derived survival and physical-condition scores had the highest impact on model predictions, followed by other LLM-derived PCIs. Notably, mobility impairment, abnormal examination findings, and B-symptoms outweighed metastatic sites in predictive importance.

**Figure 5:**
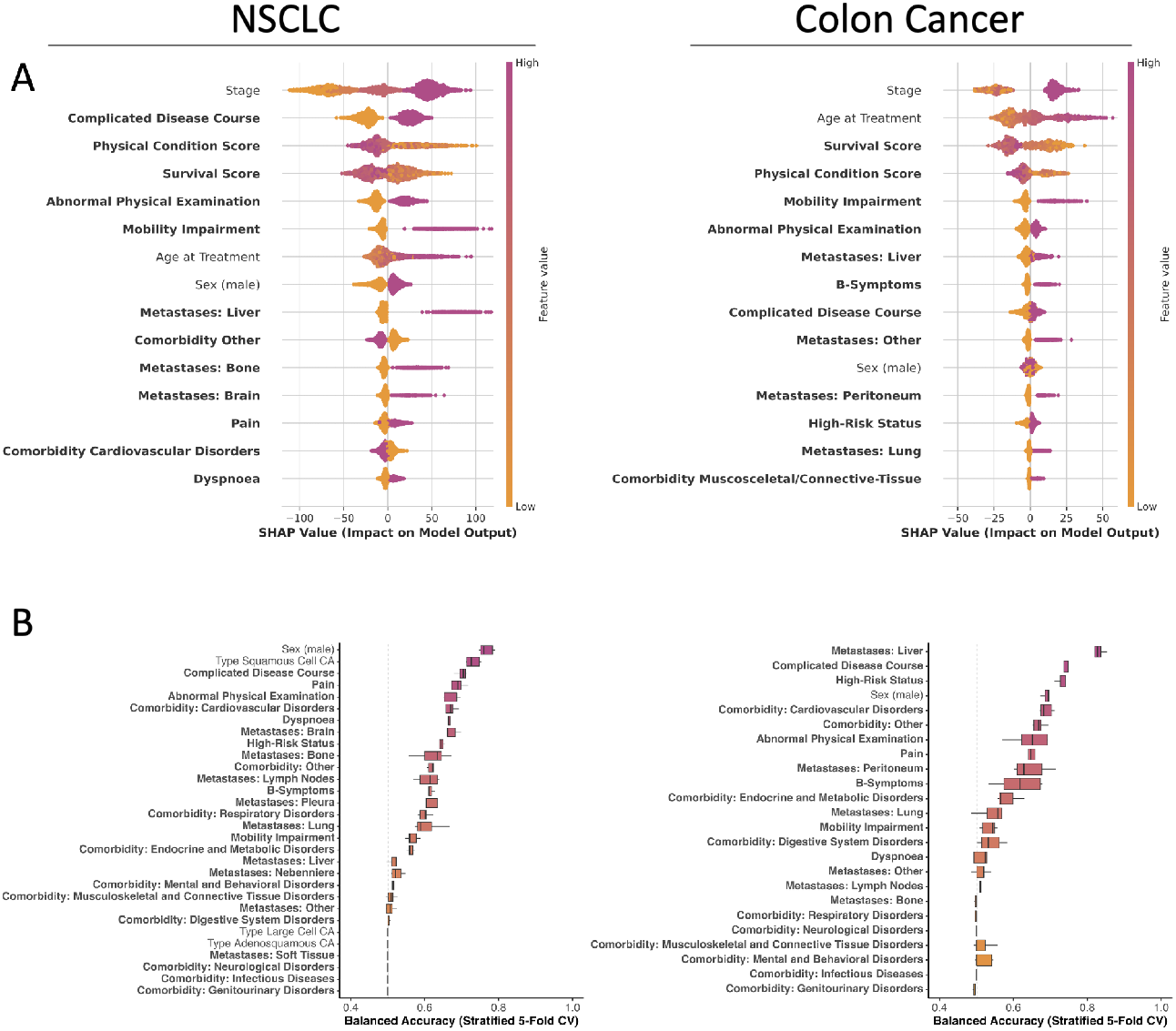
Interpreting model behavior and embedding informativeness. **A:** SHAP summary plots for the top 15 predictors in a Random Survival Forest trained on baseline clinical data plus LLM-extracted features. Features are ordered by their mean absolute SHAP values, quantifying their relative contribution to the predicted survival risk. SHAP distributions are shown over 10 cross-validation test folds. The x-axis represents risk contribution (positive = higher risk), with color indicating feature value (orange=low, purple=high). **B:** Balanced accuracy (stratified 5-fold CV) of logistic regression models trained on text embeddings from EHR notes and discharge reports to predict clinical features (LLM-extracted variables in bold). The dashed vertical line indicates chance-level balanced accuracy (0.5).

### Text embeddings encode patient characteristics

Given the robust performance of the embedding-enhanced prediction model, we investigated the information captured within the text embeddings. As the embedded features are not directly interpretable, we used a surrogate approach using regression models based on text embeddings to predict structured, understandable features (**Fig. 5B**).

In the NSCLC cohort, sex was predicted most accurately (median balanced accuracy = 0.76), followed by squamous cell carcinoma (0.73). Among LLM-derived features, complicated disease course ranked third overall (0.7), followed by pain (0.69), abnormal physical examination (0.69), cardiovascular disorders (0.67), and dyspnoea (0.66). All PCIs achieved median balanced accuracies above 0.56, indicating that these clinical features are at least partially encoded in the text-embedding model.

In the colon cancer cohort, despite generally wider interquartile ranges, the results were largely consistent with those observed in the NSCLC cohort (Spearman’s ρ=0.65, p<0.001). Liver metastases were the most accurately predicted variable in the colon cancer cohort (balanced accuracy = 0.83), followed by complicated disease course (0.75), high-risk status (0.73), and sex (0.69). Notably, dyspnoea was less predictable in this cohort (0.52), likely reflecting the lower prevalence outside of lung cancer.

## Discussion

Multimodal real-world data have become increasingly accessible across all domains and have been used in combination with deep learning models to predict outcomes and guide treatment decisions.^20,21^ In addition, a significant proportion of information in EHR consists of unstructured text that remains largely unused by current machine learning approaches.^2,22,23^ In this study, we investigated the potential of LLMs for the prediction of real-world patient outcomes based on a large dataset containing pre-treatment EHR notes and discharge summaries from 2,708 patients with NSCLC and a validation cohort of 814 patients with colon cancer.

We implemented two strategies to leverage clinical texts for patient survival prediction. First, we used an LLM for zero-shot information extraction. This resulted in seven patient condition indicators, along with each patient’s comorbidities and sites of metastasis. Comparing the LLM-extracted features with manually curated features showed high similarity, aligning with previous studies, showing the potential of LLM for accurate information extraction.^24–26^ To investigate the LLM’s intrinsic medical reasoning capabilities in a minimally guided setting, we further prompted it to generate two composite scores at the patient level: an overall physical status score and a survival score. Notably, we found that both scores have high prognostic value in our real-world cohorts, showing the remarkable medical zero-shot capabilities of current LLM without task-specific training. These results align with recent studies demonstrating that LLMs can infer clinical risk based on patient context, suggesting their potential for general-purpose clinical reasoning.^27–29^ Second, we used a text embedding model to transform raw text into high-dimensional feature vectors, a technique previously shown to improve patient outcome prediction.^15,30,31^

Comparing models trained on LLM-derived or embedding data, we found that in both approaches, incorporating information from unstructured clinical text significantly improved survival prediction compared to UICC staging alone, with an LLM-informed model achieving the highest performance overall. This enabled the reclassification of a significant proportion of NSCLC and colon cancer patients into four distinct LLM-informed risk groups. By investigating the decision-making of our model, we found that while TNM staging remained the most important feature in both NSCLC and colon cancer cohorts, the two LLM-derived composite scores consistently ranked among the top four most important features. This suggests that the LLM-derived host-level characteristics play a modulatory role, contextualizing tumor-specific information. These findings align with growing evidence that integrating host-specific factors alongside tumor-specific characteristics is critical for optimizing personalized treatment approaches in cancer care.^32–35^ Although the black-box nature of text embeddings limits interpretability, our analyses indicate that they also captured host-specific information, potentially contributing to the robust performance of embedding-enhanced survival models in our study.

Our study has limitations, most of which are inherent to the nature of retrospective real-world studies. The heterogeneity in oncology documentation, including variability in note frequency and detail, introduces potential biases and distribution shifts. To capture a representative patient population of a large academic cancer center, we included all patients at the time of their first treatment at our institution. However, some patients may have received previous external treatments, which could have influenced LLM decision-making. While this adds variability, it also reflects the realities of routine clinical care. Another limitation is inherent biases from the LLM itself, reflecting possible inaccuracies or outdated information from training datasets, such as obsolete ICD definitions. We evaluated the reliability of LLM-based information extraction against the structured EHR data of our institution. However, its reliability in diverse hospital systems and international healthcare settings remains an important area of ongoing research.

Our results show the potential of unstructured text to significantly enhance survival prediction and treatment guidance. While embedding models allow straightforward use of unstructured text, LLMs can extract interpretable information at scale, transforming fragmented narratives into actionable features that further enhance prediction models. Recent advances in open-source and distilled LLMs make it increasingly practical to embed these tools directly into clinical workflows. Looking ahead, the integration of LLMs into real-world EHR systems could transform precision medicine by using unstructured text data to develop personalized treatment strategies.

## Methods

### Study design

We retrospectively included 2,708 non-small cell lung cancer (NSCLC) patients and 814 colon cancer patients treated at University Hospital Essen in this study. Overall survival (OS) was defined as the time from initiation of treatment at our institution to death from any cause. The date of death was obtained from either the medical record or, if unavailable, from the state cancer registry. Patients without a documented date of death were right censored on the date of their last clinical visit. The study was approved by the Ethics Committee of the Medical Faculty of the University Duisburg-Essen (No. 22-10881-BO).

### Data acquisition

All medical data used in this study were collected using the Smart Hospital Information Platform (SHIP) of the University Hospital Essen. With SHIP, the data of multiple clinical subsystems are integrated and stored in Fast Healthcare Interoperability Resources (FHIR) format, allowing for data collection based on specific queries. After the collection of NSCLC and colon cancer patients based on available tumor documentation, we excluded patients without text reports before the start of cancer treatment at our institution. For the resulting patient cohort, further characteristics, such as demographics and ICD-10 codes, were queried. The entire data collection process is shown in **Supp. Fig. 5**.

### Text processing and model deployment

We used two approaches to incorporate information from unstructured clinical text into our analyses:

#### 1. LLM Feature Extraction

We used Meta’s multimodal large language model, Llama 4 Scout, to extract pre-specified features in this study based on a uniform prompt in a zero-shot manner (**Supp. Tab. 4**). The Scout model, a version of the Llama 4 series, features 17 billion active parameters, with a total of 109 billion parameters distributed across 16 experts. Llama 4 Scout has been deployed locally using Hugging Face’s Text Generation Interference (TGI) on four NVIDIA H100 GPUs, operating with a context window of 8,000 tokens. In particular, we extracted metastasis locations and comorbidities as lists, representing comorbidities by ICD codes. Furthermore, we encoded the presence or absence of B-symptoms, dyspnoea, pain, high-risk status, complicated disease course, abnormal physical examination, and mobility impairment as binary features. Beyond these, we prompted the LLM to generate two composite scores based on the reports: a Physical Condition Score (ranging from 0 to 100, where scores closer to 0 indicate worse physical condition) and a Survival Score (ranging from 0 to 100, where 100 indicates a higher likelihood of survival). Outputs were given in JSON format, and cases with failed extraction were excluded from subsequent analyses (**Supp. Fig. 5**).

#### 2. Text embeddings

To generate high-dimensional vector representations directly from the raw clinical text, we used the multi-lingual BGE-M3 model, which is capable of handling inputs of varying sizes.^15^ A maximal chunk size of 512 tokens and a sliding window size of 32 tokens was used for dense retrieval.

To ensure the confidentiality of patient data, the text reports were exclusively processed within the hospital network. All models were internally deployed using the KI Translation Essen (KITE) platform. KITE, a research infrastructure of the University Hospital Essen and the Medical Faculty of the University of Duisburg-Essen, supports the translation of algorithms to the point of care.

### Datasets

For evaluation, we created three datasets to assess whether these two approaches for information extraction/embedding can improve survival prediction as measured by the C-Index. Therefore, we created:

#### 1. Stage Dataset

This dataset includes only the tumor stage information (I-IV) of each patient.

#### 2. Baseline Dataset

The stage dataset was expanded through the incorporation of additional patient demographics and clinical variables, including sex, age, and, for patients with NSCLC, tumor type. This expanded dataset was designated as the baseline dataset. We then enriched the baseline dataset with features extracted and embeddings generated by an LLM. To maximize patient inclusion, we merged data from both EHR notes and clinical discharge reports, keeping patients with at least one report type. When only one report was available, we used that data directly and when both were present, we combined the features as follows:

- **Categorical Features:** We applied a logical OR to merge values. For example, if “pain” was marked as false in the EHR note but true in the discharge report, the combined value was set to true. Comorbidities and metastases were dummy encoded beforehand.
- **Continuous Features:** For variables such as embeddings, survival scores, and physical condition, we calculated the average value.

This process produced two distinct datasets: Baseline + LLM-extractions and Baseline + text-embeddings. For the latter, which contained high-dimensional embeddings, we applied principal component analysis on the embeddings (PCA) to distill the feature space down to the top 30 components.

### Curating LLM-Extracted Features

We identified common comorbidities using ICD codes and grouped them into clinically relevant categories. To focus on non-cancer comorbidities, we excluded ICD codes that start with “C”. We identified risk groups by ICD code chapter, Cardiovascular Disorders (I), Respiratory Disorders (J), Endocrine and Metabolic Disorders (E), Infectious Diseases (A, B, U), Neurological Disorders (G), Digestive System Disorders (K), Genitourinary Disorders (N), Musculoskeletal and Connective Tissue Disorders (M), and Mental and Behavioral Disorders (F), and assigned all remaining codes to an Other category. Only ICD codes appearing in at least 25 reports were used, and any code not fitting a predefined group was classified as “Other”. For metastases, we standardized the extracted terms, accounting for variations like plural versus singular forms and abbreviations, by mapping them to common metastases locations. We retained only those metastases occurring in at least 25 reports, applying the same grouping strategy as with ICD codes, where unmatched terms were assigned to “Other”. Absence of comorbidities was encoded by setting all comorbidity risk group flags to False, while absence of metastases was encoded separately by setting all metastasis flags to False.

### Survival Model

We trained a Random Survival Forest (RSF) with 1,000 trees, a minimum of five samples required to split an internal node, and at least five samples per leaf, using 10-fold cross-validation on the three datasets, Baseline, Baseline + LLM-extractions and Baseline + text-embeddings. To derive a five-year risk score, we set a reference time T_ref of 1825 days and evaluated each test patient’s cumulative hazard function at T_ref. That hazard value served as the patient’s risk score, which we then partitioned into four groups. To limit the influence of extreme values, we excluded scores below the 2.5th percentile and above the 97.25th percentile when calculating bin boundaries, performed equal-width binning on the remaining range, and assigned all excluded scores to the lowest or highest bin as appropriate.

### Statistics

We preprocessed the 1,024-dimensional text embeddings of discharge reports and EHR notes via principal component analysis (PCA) in Python’s *scikit-learn* package to reduce dimensionality to 30 features using default settings.^36^ Continuous features were standardized using *scikit-learn’s* StandardScaler, and categorical features were one-hot encoded with its MultiLabelBinarizer. Kaplan-Meier curves were estimated and compared by the log-rank test, and Cox proportional hazards models were fitted using the *survival* package in R.^37,38^ Random Survival Forests were trained independently on each dataset using tenfold cross-validation in Python’s *scikit-survival* package to estimate C-Indices, which were then compared across datasets by paired Wilcoxon signed-rank test in R’s *stats* package.^39^ SHAP values were calculated with the model-agnostic explainer from the Python *shap* library for each fold and aggregated into a single explainer object, enabling consistent, global interpretation of feature contributions across the entire dataset.^19^ Spearman correlations of SHAP values were computed using Python’s *scipy* package, and Pearson correlations between LLM-inferred scores and patient characteristics were calculated with R’s *stats* package.^40^

## Data availability

Anonymized data are available from the corresponding author upon reasonable request. Data cannot be shared with investigators outside the institution without consent.

## Acknowledgements

The data for this project was provided by the Smart Hospital Information Platform (SHIP), managed by the Data Integration Center at the University Medicine Essen. SHIP serves as a comprehensive digital health platform for integrating data from all major clinical subsystems using a holistic FHIR-based approach. It enables the purification, analysis, distribution, and visualization of clinical data.

J.T.S. is grateful for support from the German Cancer Consortium (DKTK).

## Author contributions

Conceptualization: JKeyl, JKleesiek, PK

Methodology: NK, TL, MP, GM, PK, JKleesiek, Jkeyl

Formal analysis: NK, TL, PK, JKeyl

Investigation: all authors

Data acquisition and evaluation: JF, SK, EK, JE, MP, AJ, JTS, MW, SK, UPN, FK, SH, MS, PK, JKleesiek, JKeyl

Data curation: TL, JKeyl

Writing - original draft: NK, TL, AD, PK, JKleesiek, Jkeyl

Writing - review and editing: all authors

Supervision: JKeyl, JKleesiek, PK

