## Supplementary Material for "Large Language Models Improve Cancer Survival Prediction Using Real-World Clinical Notes"

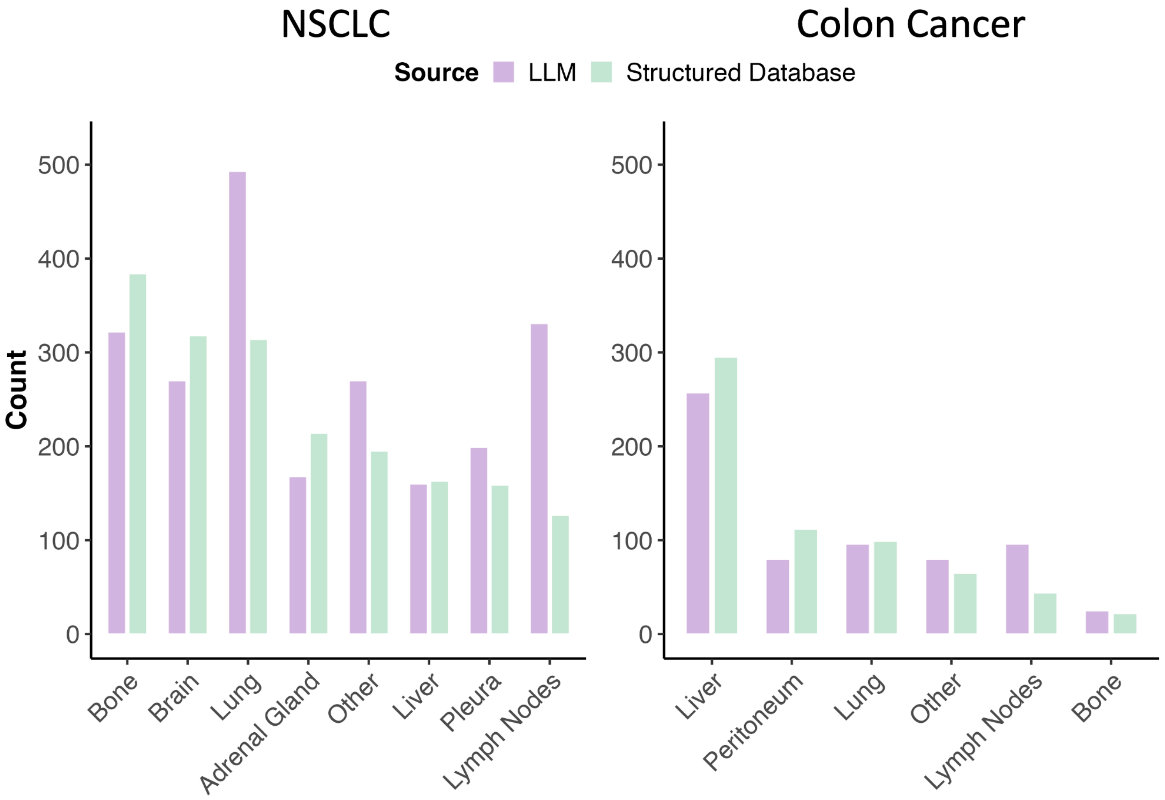


**Supplementary Figure 1: Comparison of metastatic site frequencies in stage IV NSCLC (left) and colon cancer (right) between a structured clinical database (green) and our LLM-based extraction from the unstructured medical text records (purple).** Each bar shows the number of patients with metastases at a given anatomical location, sites that together account for less than 3 % of all metastasis occurrences across both sources (threshold = total counts × 0.03) were pooled into the "Other" category.

**Supplementary Table 1: Results of univariate and multivariate Cox proportional-hazards models for colon cancer cohort evaluating the association between LLM-extracted covariates and overall survival.** In the univariate analysis, each feature was tested individually. For each covariate, the hazard ratio (HR), 95 % confidence interval (CI) and p-value are reported. Structured EHR data comprises all fields originally available in structured format, whereas LLM-inferred variables are those derived by the model from unstructured medical documentation.

| | **Univariable analysis** | | **Multivariable analysis** | |
|---|---|---|---|---|
| **Structured EHR Data** | **HR (95% CI)** | **P value** | **HR (95% CI)** | **P value** |
| Age at Treatment (per 1 SD) | 1.24 (1.1-1.4) | **<0.001** | 1.44 (1.26-1.64) | **<0.001** |
| Stage II vs I | 0.62 (0.31-1.22) | 0.163 | 0.61 (0.31-1.21) | 0.160 |
| Stage III vs I | 0.84 (0.45-1.55) | 0.570 | 0.92 (0.49-1.71) | 0.786 |
| Stage IV vs I | 2.61 (1.57-4.33) | **<0.001** | 2.84 (1.67-4.85) | **<0.001** |
| Sex (male) | 0.88 (0.7-1.11) | 0.278 | 0.93 (0.74-1.17) | 0.552 |
| **LLM-Inferred Variables** | | | | |
| High-Risk Status | 1.85 (1.42-2.42) | **<0.001** | 1.01 (0.71-1.45) | 0.935 |
| Abnormal Physical Examination | 1.79 (1.43-2.25) | **<0.001** | 1.27 (0.96-1.69) | 0.100 |
| Dyspnoea | 1.61 (1.14-2.27) | **0.007** | 1.12 (0.78-1.62) | 0.529 |
| Complicated Disease Course | 1.92 (1.5-2.45) | **<0.001** | 1.12 (0.8-1.57) | 0.518 |
| B-Symptoms | 2.11 (1.63-2.74) | **<0.001** | 1.4 (1.04-1.89) | **0.025** |
| Pain | 1.28 (1.02-1.61) | **0.031** | 0.98 (0.76-1.26) | 0.879 |
| Mobility Impairment | 2.4 (1.8-3.19) | **<0.001** | 1.79 (1.3-2.47) | **<0.001** |

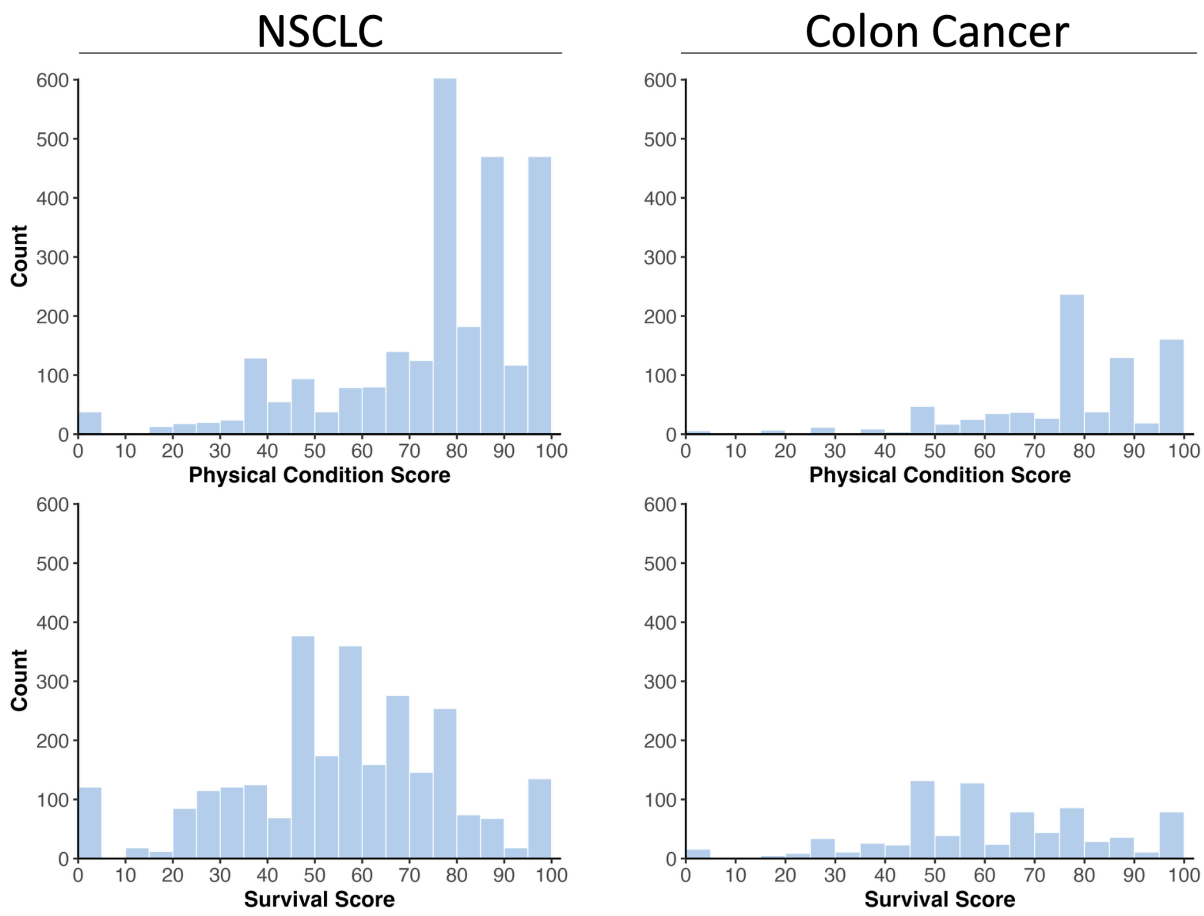


**Supplementary Figure 2: Histogram distributions of LLM-created composite scores for physical condition and survival in NSCLC and colon cancer cohorts.**

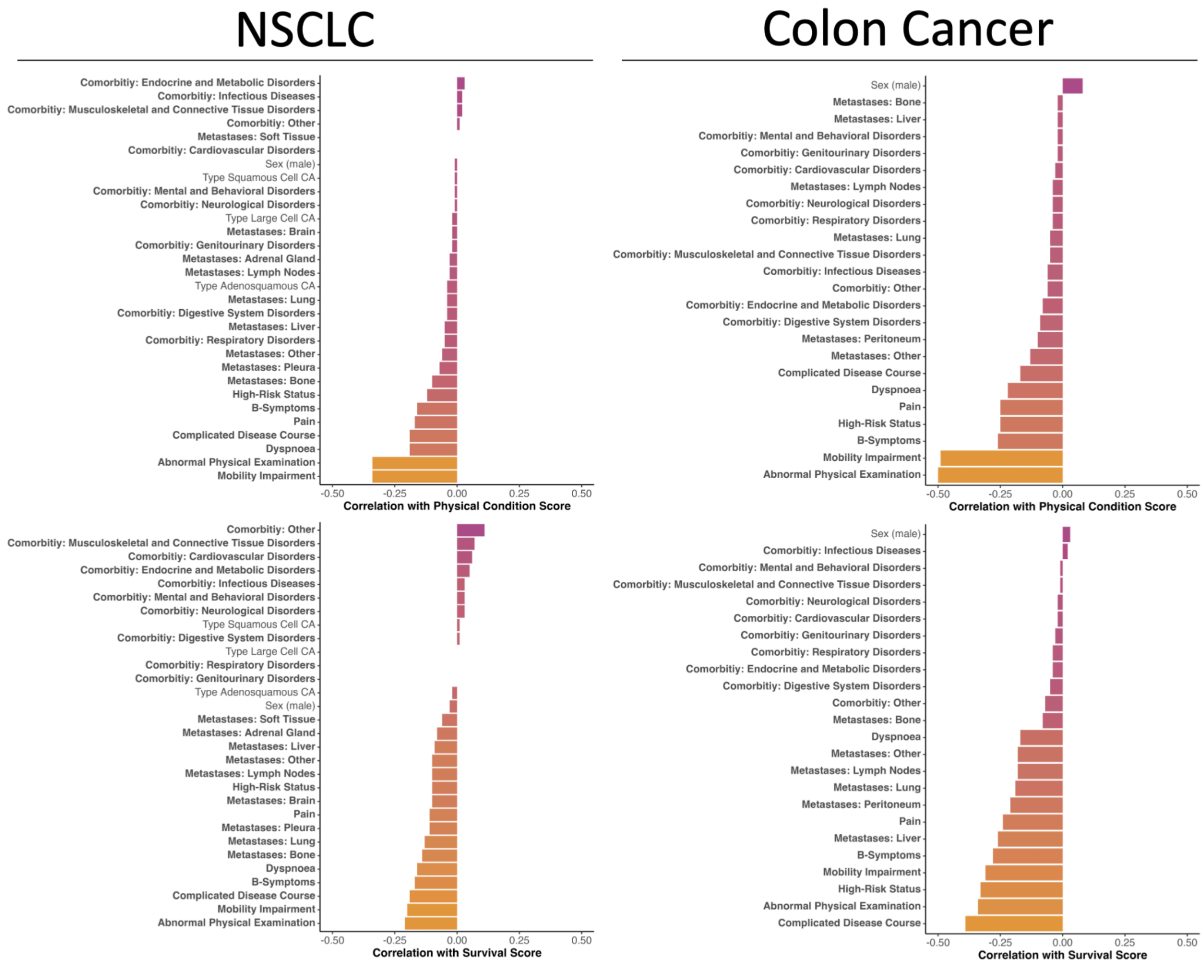


**Supplementary Figure 3: Association of LLM-inferred scores with patient characteristics.** Pearson correlations between each binary feature and the two continuous scores: physical condition (top row) and survival (bottom row) for the NSCLC (left) and colon cancer (right) cohorts. Feature names highlighted in bold were extracted by the LLM.

**Supplementary Table 2: Results of univariate and multivariate Cox proportional-hazards models for NSCLC cohort evaluating the association between LLM-inferred scores and overall survival.** In the univariate analysis, each feature was tested individually. For each covariate, the hazard ratio (HR), 95 % confidence interval (CI) and p-value are reported. Structured EHR data comprises all fields originally available in structured format, whereas LLM-inferred variables are those derived by the model from unstructured medical documentation.

| | **Univariable analysis** | | **Multivariable analysis** | |
|---|---|---|---|---|
| **Structured EHR Data** | **HR (95% CI)** | **P value** | **HR (95% CI)** | **P value** |
| Age at Treatment (per 1 SD) | 1.03 (0.97-1.1) | 0.301 | 1.1 (1.03-1.17) | **0.004** |
| Stage II vs I | 1.53 (1.08-2.15) | **0.016** | 1.43 (1.01-2.02) | **0.042** |
| Stage III vs I | 2.34 (1.8-3.04) | **<0.001** | 2.47 (1.9-3.21) | **<0.001** |
| Stage IV vs I | 4.1 (3.26-5.14) | **<0.001** | 4.51 (3.57-5.69) | **<0.001** |
| Sex (male) | 1.34 (1.18-1.53) | **<0.001** | 1.33 (1.17-1.52) | **<0.001** |
| **LLM-Inferred Variables** | | | | |
| Physical Condition Score (per 1 SD) | 0.82 (0.78-0.86) | **<0.001** | 0.79 (0.74-0.84) | **<0.001** |
| Survival Score (per 1 SD) | 0.81 (0.77-0.86) | **<0.001** | 0.97 (0.9-1.05) | 0.471 |

**Supplementary Table 3: Results of univariate and multivariate Cox proportional-hazards models for colon cancer cohort evaluating the association between LLM-inferred scores and overall survival.** In the univariate analysis, each feature was tested individually. For each covariate, the hazard ratio (HR), 95 % confidence interval (CI) and p-value are reported. Structured EHR data comprises all fields originally available in structured format, whereas LLM-inferred variables are those derived by the model from unstructured medical documentation.

| | **Univariable analysis** | | **Multivariable analysis** | |
|---|---|---|---|---|
| **Structured EHR Data** | **HR (95% CI)** | **P value** | **HR (95% CI)** | **P value** |
| Age at Treatment (per 1 SD) | 1.24 (1.1-1.4) | **<0.001** | 1.42 (1.24-1.62) | **<0.001** |
| Stage II vs I | 0.62 (0.31-1.22) | 0.163 | 0.52 (0.26-1.04) | 0.066 |
| Stage III vs I | 0.84 (0.45-1.55) | 0.570 | 0.78 (0.42-1.47) | 0.448 |
| Stage IV vs I | 2.61 (1.57-4.33) | **<0.001** | 2.71 (1.6-4.58) | **<0.001** |
| Sex (male) | 0.88 (0.7-1.11) | 0.278 | 0.87 (0.69-1.09) | 0.234 |
| **LLM-Inferred Variables** | | | | |
| Physical Condition Score (per 1 SD) | 0.79 (0.72-0.86) | **<0.001** | 0.86 (0.75-0.98) | **0.021** |
| Survival Score (per 1 SD) | 0.69 (0.62-0.77) | **<0.001** | 0.81 (0.7-0.95) | **0.010** |

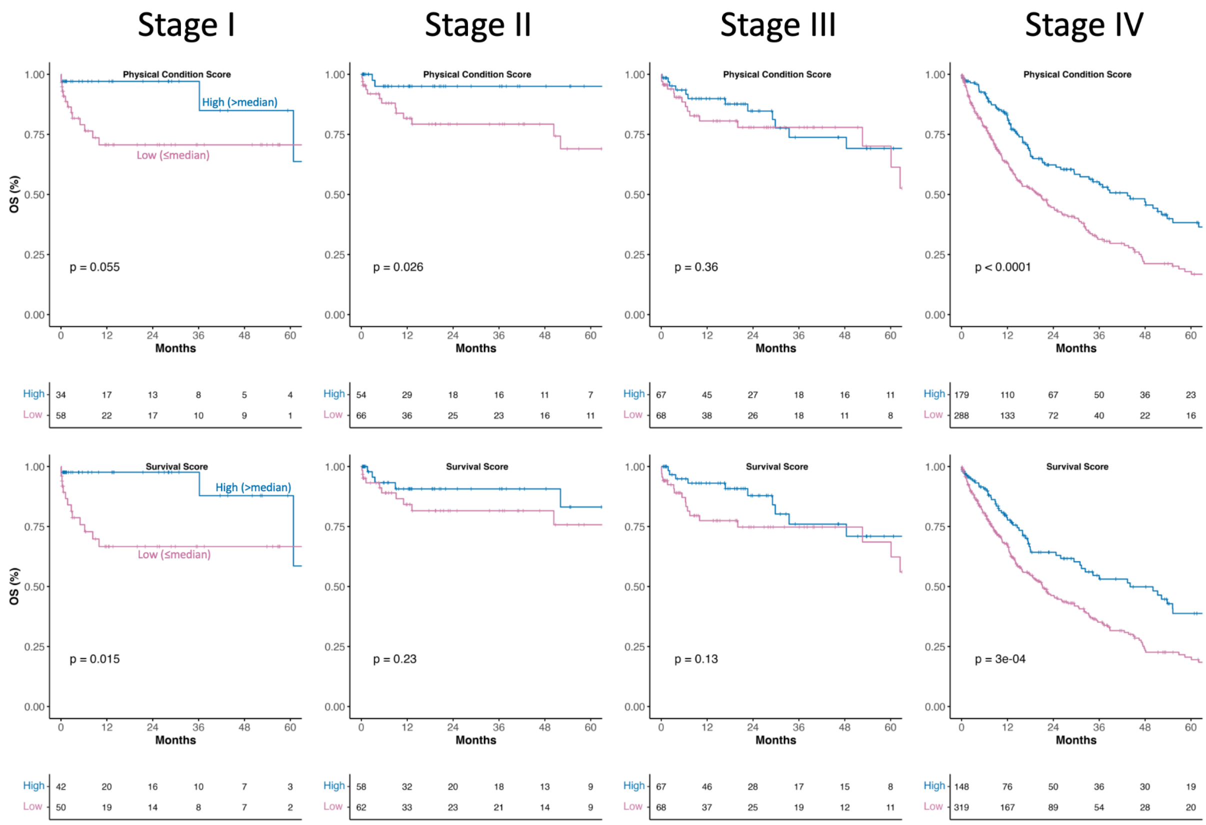


**Supplementary Figure 4: Kaplan-Meier survival curves for colon cancer patients stratified by LLM-inferred physical condition and survival scores at disease stages I–IV.** Patients were dichotomized by the median within their respective stage into high-score (> median; blue) and low-score (≤ median; magenta) groups. Survival differences between groups were assessed by the log-rank test, and corresponding p-values are indicated in each panel.

**Supplementary Table 4: Base prompt for LLM-driven information retrieval from clinical notes.**

| |
|---|
| You are a clinical expert in oncology. Extract all the information mentioned in the following example or deduce it logically. Important instructions:<br>– The metastasis location must contain a list from the specified categories. If none of the specific categories are mentioned, return “Other” or “Unknown.”<br>– If multiple locations are mentioned, extract all of them as a list.<br>– Make sure that for “Physical condition,” a score is calculated for each patient that reflects their general physical condition (the higher the score, the better the general condition).<br>– Make sure that for “Survival score,” a score is calculated for each patient that reflects their probable survival time (the higher the score, the more favorable the prognosis).<br>– Make sure that the JSON format is valid and that no information is outside of “” or without formatting. Do not include additional explanations in the JSON output for the ICD-10 codes.<br><br>Extract only the information mentioned in the example. Return the results in validly formatted JSON format, exactly as in the following example:<br>{…} |

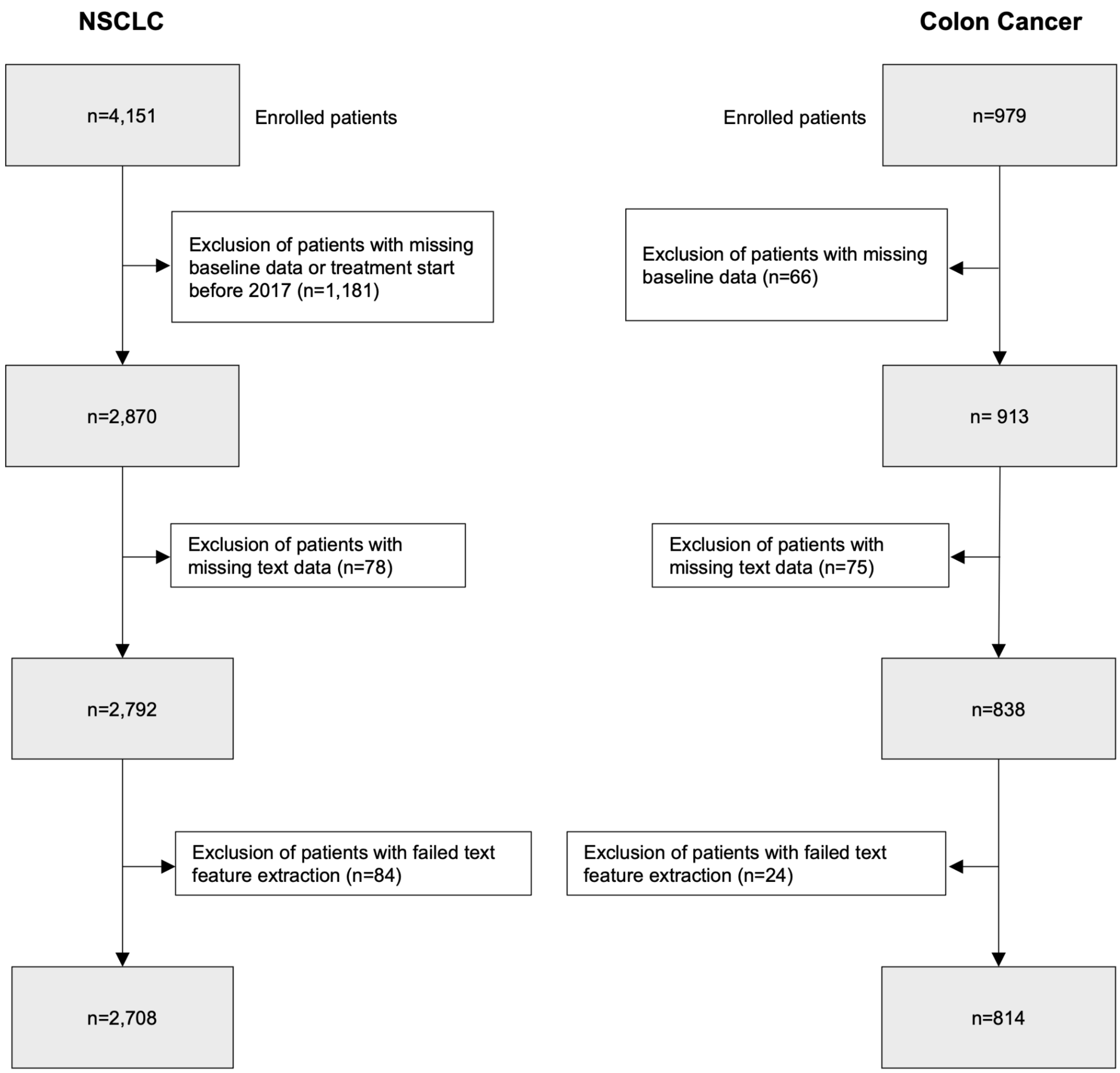


**Supplementary Figure 5: Flow diagram for patients included in the analyses.**
